# Economic Impact of a Large-Scale Scabies Upsurge on Healthcare Facilities in Rohingya Refugee Camps: A Retrospective Costing Study

**DOI:** 10.64898/2025.12.18.25342638

**Authors:** Charls Erik Halder, Md Abeed Hasan, James Charles Okello, Sayed Sunny Uz Zaman, Julekha Tabassum Poly, Hamim Tassdik, Dickson Wafula Barasa, Emmanuel Roba Soma, Md Farhad Hussain, U Maung Prue, Sirajul Munir Khandokar, John Patrick Almedia, Jahangir Alam

## Abstract

**Background:** Scabies is one of the common infectious skin conditions globally, with a significantly high burden in hot and tropical countries, resource-poor settings, and areas with high population density. Rohingya refugee camps in Cox’s Bazar are one of the most protracted refugee crises in the world, sheltering approximately 1,143,096 refugees in 33 overcrowded refugee camps. While most existing literature focuses on mass drug administration (MDA) interventions or community-level estimates, the economic burden of scabies on health system is rarely studied and documented.

**Methods:** This is a retrospective cost study, where we have used financial and epidemiological data from January 2021 to December 2024. Costing was done from the provider’s perspective, focused on what International Organization for Migration (IOM) spent during the period as a health service provider. Acombination of standard stepdown approach and micro-costing methods were used. Financial data for the study period 2021 to 2024 were collected from the IOM health programme’s annual budget and consumption reports. The study population included all individuals who were clinically diagnosed with scabies and received care at 35 IOM-supported health facilities in Ukhiya and Teknaf, Bangladesh.

**Results:** The overall estimated financial cost for IOM’s scabies outbreak response through its health facilities was USD 2.12 million, with an annual average of USD 531,729. The average cost per scabies management ranged between USD 5.33 and USD 6.54, with the highest in 2021 and the lowest in 2022. Drug costs accounted for 11.92% of the overall cost over 4 years. Of the total cost of $253,629.43 over 4 years, 79% was attributed to permethrin topical cream, which was used to manage an estimated 85% of the total managed cases. Scenario analysis demonstrates that the existing permethrin-based treatment preference is the most expensive treatment modality, compared with ivermectin-based treatment and mixed-treatment approaches.

**Conclusion:** Although the average cost of treating scabies is relatively low, overall, the treatment cost for such a large population has a significant economic impact. This study found a substantial effect of mass drug administration on reducing the financial burden on the healthcare system.

## INTRODUCTION

Scabies is one of the common skin conditions globally with significantly high burden in hot and tropical countries, resource poor settings and in areas with high population density. According to World Health Organization, the global incidence of scabies at any given time is more than 200 million, affecting 5 to 50 per cent of children in resource-poor areas (1). Refugees and the displaced population possess a higher risk of scabies outbreak due to overcrowded living arrangements, insufficient access to water, sanitation, and laundering facility, limited access to healthcare services and shortage of essential medicines (e.g. scabicides) (2,3).

Scabies is a curable condition, effectively treatable with the topical application of permethrin or oral administration of ivermectin. Unlike non-communicable diseases or high burden communicable diseases, such as, Tuberculosis, HIV and Hepatitis, uncomplicated scabies does not require long-term treatment. The treatment cost of scabies, if considered as single unit, is relatively inexpensive. However, scabies has the high potentiality to cause massive outbreak, especially in overcrowded and refugee settings. Overlooking its collective economic burden in an outbreak setting may create misperception among the policy makers and public health professionals regarding its potential economic impact and the need for adequate planning of resources. For instance, an study in Fiji reported that although the average cost per presentation for scabies at primary healthcare was US$17.7, the estimated annual healthcare costs of scabies and its complications was US$3.0 million equivalent to per capita cost of US$3.3 (4).

The Rohingya refugee camps in Cox’s Bazar is one of the largest protracted refugee crises in the world sheltering approximately, 1,143,096 refugees in 33 overcrowded refugee camps (5). Overcrowded living arrangements, fragile shelters, inadequate water, sanitation, and hygiene (WASH) facilities together with heavy monsoons in both the refugee camps and neighboring host communities often result in outbreak of infectious diseases, diphtheria, measles, COVID-19, and Acute Watery Diarrhea (AWD)/cholera. In the first study of our research series on scabies in this setting, we reported a massive outbreak that occurred between 2021 and 2024, with a total of 384,852 reported cases with an overall attack rate of 5,562.59 cases per 10,000 population over the four-year period *(Halder et. al)* The high burden of scabies has led to an increased demand for heightened medical attention and additional resources. Simultaneously, it contributed to disruptions in daily life, potentially hindering social interactions and economic activities within the refugee community.

The disease burden of scabies infestation, along with its associated complications, places a significant financial burden on healthcare systems. It not only impacts the costs associated with medicines (e.g., topical scabicides, antibiotics for secondary infections) and diagnostics but a huge strain over spacing, infection prevention and control, health workforce and quality of service exhausting the available resources of an already overstrained health system (6).

Scabies also can lead to secondary bacterial infection, which could be complicated to life-threatening conditions, like acute glomerulonephritis and rheumatic heart fever, further efforts of healthcare workers and cost of the health system for managing the complications (7). While most existing literature focuses on mass drug administration (MDA) interventions or community-level estimates, economic burden of scabies on health facility is rarely studied and documented (4,8). However, understanding the direct healthcare costs of scabies is critical for effective planning of intervention, resource allocation and initializing cost-effective solutions, like MDA. To address this research gap, this study evaluated the financial impact of the massive upsurge of scabies on healthcare system. This research will assist the health programme and policy makers to understand financial implication of scabies upsurge and thus, facilitate proper planning for effective integration of scabies prevention and management within the essential health service model. This will also assist to examine potential impact and cost effectiveness of new prevention and management strategies or programme.

## METHODOLOGY

### STUDY SETTING AND POPULATION

The study implemented in Rohingya refugee camps in Ukhiya and Teknaf in Cox’s Bazar. The study population included all individuals who were clinically diagnosed with scabies and received care at 35 IOM-supported health facilities in Ukhiya and Teknaf, Bangladesh, between January 1, 2021 – April 31, 2024. Scabies caused a massive upsurge during this period in the refugee camps that prompted a substantial response from IOM. Therefore, these camps were selected for this study to get an in-depth information of the outbreak.

All scabies cases were diagnosed clinically at 35 IOM supported facilities in Ukhiya and Teknaf. MSF clinical guidelines – diagnosis and treatment manual was used for diagnosis and clinical management of the cases. 5% permethrin cream was mainly used for treatment of scabies.

### STUDY DESIGN

This is a retrospective costing study, where we have used financial and epidemiological data from January 2021 to December 2024. Costing was done from provider perspective focused on what IOM spent during the period as health service provider. A combination of standard stepdown accounting and micro-costing methods were used (9,10).

Micro-costing (or ingredient-based costing) was calculated direct cost per-case by itemizing resources used per patient (11). In this study, we estimated the cost per patient for medicines required, laboratory cost, and supplies for infection prevention and control.

Step-down costing method was used to estimate costing related to capitals, coordination, supervision, operation and maintenance, capacity building, risk communication and community engagement, volunteer payment and salaries and benefits of healthcare workers, meaning that total costs and resources utilized by the whole health programme of IOM during the period was systematically and proportionately distributed to the “specific service unit”. The specific service unit refers, hereby, the scabies consultation and treatment service (12).

The study did not take account of the economic costs (value of resources that could have been productively used elsewhere) such as the value of volunteer time, donated goods, shared infrastructure in the scabies management. All staff and volunteers involved were salaried under the program, and all supplies, equipment, and logistics were directly financed by IOM.

### DATA COLLECTION AND MANAGEMENT

Financial data for the study period 2021 to 2024 were collected from the IOM health programme’s annual budget and consumption reports. A cost recording and data analysis tool was developed to record and analyze all relevant costs associated with scabies response (Supplementary 1). Field visits to health facilities and meeting with healthcare workers and medical logisticians were made to develop the cost recording and data analysis tool. All costs were categorized by a) capital investment and b) recurrent costs.

For shared resources, for instance, human resources (healthcare workers and volunteers), coordination, operations and maintenance, and laboratory costs, annual costs were estimated based on the total expenditure reported in the financial documents and relevant procurement orders of the programme. The total costs were then proportionally allocated for scabies using the morbidity burden of scabies in comparison to the total outpatient consultations (Table 1). Scabies response specific investment includes capacity building of healthcare workers and community health workers and development and printing of information, education and communication materials.

**Table 1.** Year-wise case load and proportional morbidity of scabies.

|  | 2021 | 2022 | 2023 | 2024 |
| --- | --- | --- | --- | --- |
| Scabies | 32,046 | 140,590 | 150,470 | 61,746 |
| Infected scabies | 2,083 | 7,348 | 10,577 | 2,312 |
| Proportional morbidity (over total consultation) | 0.04 | 0.11 | 0.12 | 0.05 |

**Table 2.**
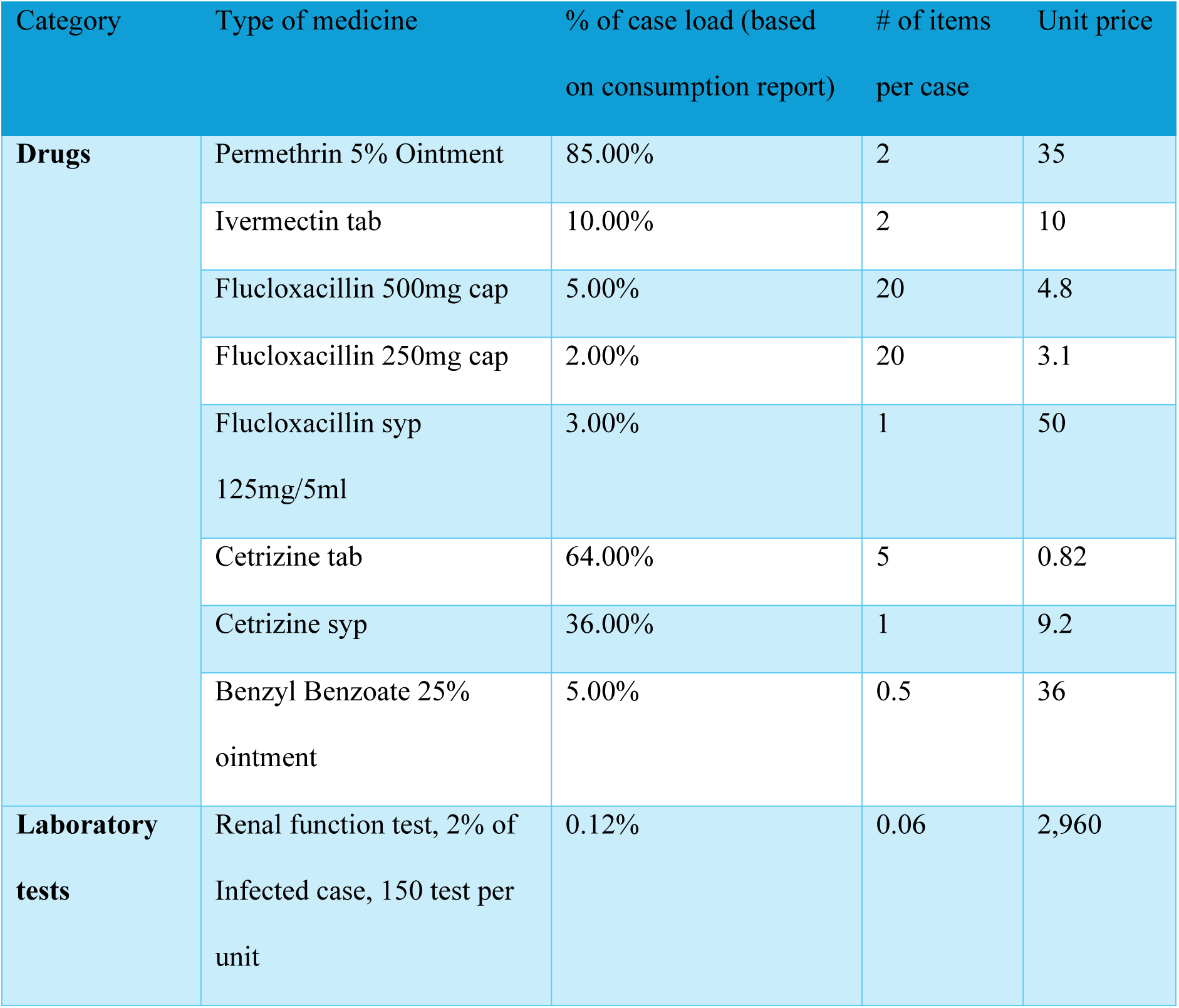
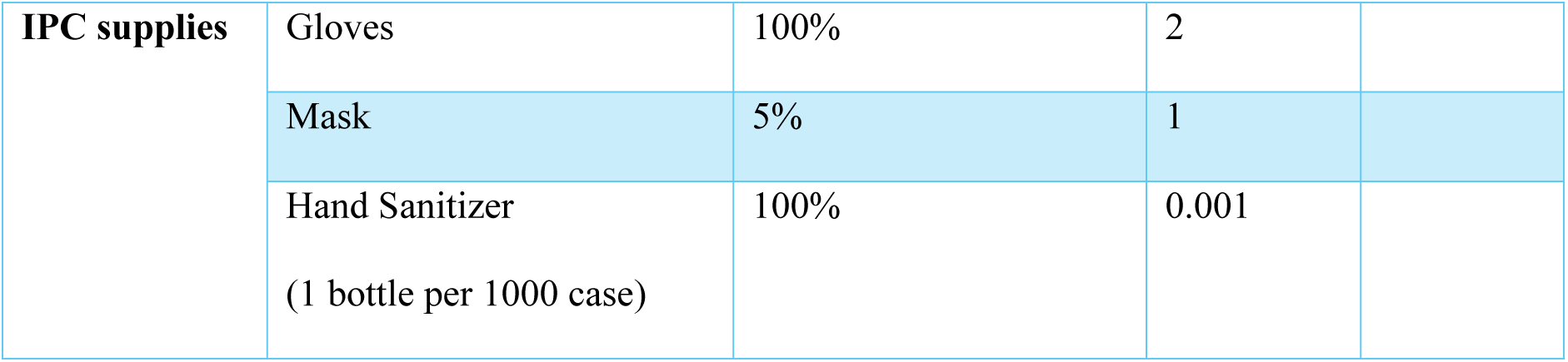
Basis and assumptions of calculating costing of drugs and laboratory tests.

Capital costs included biomedical and clinical equipment used for scabies patient management, and costs related to health facility construction/renovation. The costs were annualized using the formula below, assuming the lifespan of construction for 20 years and clinical equipment for 3 years (Creese and Parker, 1994; MSF).

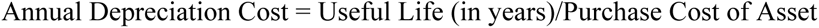

Coordination costs include salaries of programme staff and office costs at Cox’s Bazar and IOM standard 7% overhead cost over the total operation and staff cost. Supervision cost includes salaries of supervisory staff, e.g. clinical supervisors, health facility in-charges and national officers.

The cost of drugs was calculated on per case basis by considering the number of drugs given per patient according to standard clinical protocol and procurement prices as specified in the procurement order. The below assumption was made to determine the drug and laboratory cost. The percentage of case load utilizing a specific medicine was determined by reviewing the consumption report. A 3% discounting rate was used to factor for inflation across year, aligning with WHO guidance (13).

Operation and maintenance costs for health facilities included repair and maintenance, rental of land and spaces, generator and utilities, fuel, vehicle maintenance and security cost. Risk Communication and Community Engagement (RCCE) include costing for organizing awareness raising events, focus group discussions, courtyard sessions, and information, education and communication materials dedicated for scabies prevention and management. Salaries and benefits are counted for healthcare workers engaged in providing outpatient services, including triage, consultation, nursing and medication/pharmacy. Remuneration was costed for community health workers (CHW) engaged in RCCE and facility-based volunteers engaged in triage, counselling and crowd control.

#### Scenario analyses

A scenario-based costing analysis was conducted to measure the impact of different choice of treatment on the costing of scabies management at health facilities to find out the best approach for cost-efficient treatment of scabies. Scenarios were developed by the public health experts of IOM and based on different proportions of supply availability of permethrin, ivermectin and benzyl benzoate (Table 3). All other capital and recurrent costs were held constant to isolate the effect of different level of supply availability on per-case patient costing.

**Table 3.** Different planning scenario of cost analyses based on different proportion of supply availability of scabicidal agents.

|  | Scenario I<br>(Current) | Scenario II | Scenario III |
| --- | --- | --- | --- |
|  | Permethrin-based | Ivermectin-based | Equal permethrin & Ivermectin |
| **Permethrin** | 85% | 15%* | 47.5% |
| **Ivermectin** | 10%** | 80% | 47.5% |
| **Benzyl Benzoate\*\*\*** | 5% | 5% | 5% |

\* Required to manage children <15 kg and pregnant women
\*\* Required to manage complicated cases
\*\*\* Kept constant throughout as back up to support in case rupture of both permethrin and ivermectin

In current practice (scenario 1), each patient receives two tubes of permethrin at their first visit in the clinic and based on discussion with clinicians, roughly two family members per household visit the clinics. Thus, a family of four members of different age groups roughly receives four permethrin tubes, which are shared across the members for two applications in one week apart. Ivermectin tablets are reserved only for patients with secondary bacterial infections.

In scenarios 2 and 3, since Ivermectin covers regular scabies cases (not only infected), to bring household contacts under treatment one patient is allocated on average 5 Ivermectin tablets of 6 mg with the same assumption that roughly two members per family accesses the health service at health facility.

### ETHICAL CONSIDERATION

We received ethical clearance from the Ethical Review Board, Cox’s Bazar Medical College Hospital (CoxMC/2023/017). Clearance was obtained from the office of the civil surgeon and the Refugee Relief and Repatriation, who were responsible for overseeing health response in the Rohingya refugee camps in Cox’s Bazar, Bangladesh. This was a retrospective observational study, and thus, only utilized historical anonymized and aggregated data from the outbreak database. All data presented in the study were gathered during the public health outbreak response; therefore, ethical clearance prior to the data collection was not obtained.

### RESULT

Table 4 demonstrates the annual cost breakdown of scabies outbreak response from the years 2021to 2024 at IOM-supported health facilities in the Rohingya refugee camps and host communities in Cox’s Bazar. The overall estimated financial cost for IOM’s scabies outbreak response through its health facilities was USD 2.12 million with an annual average of USD 531,729.

**Table 4:**
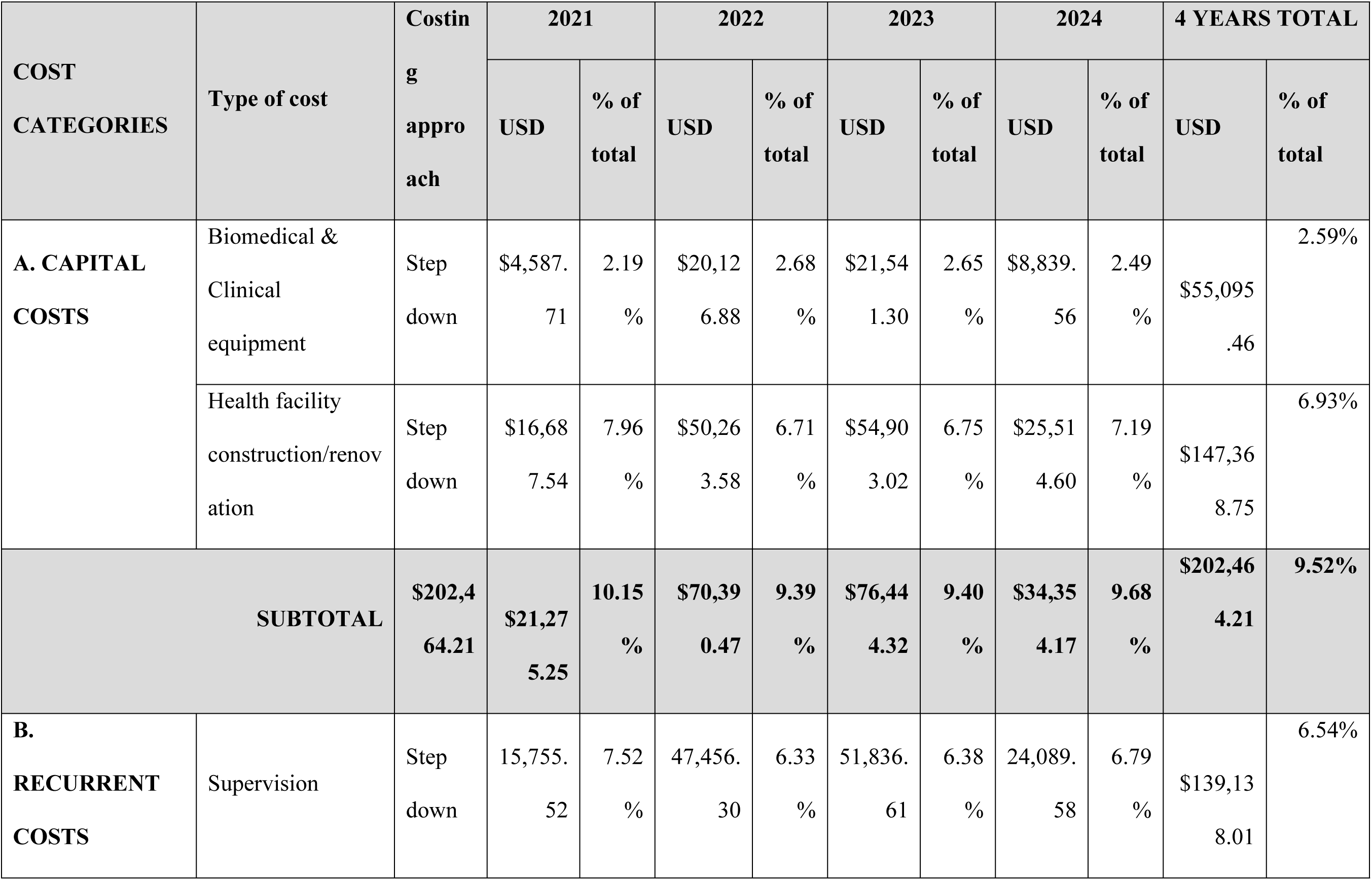
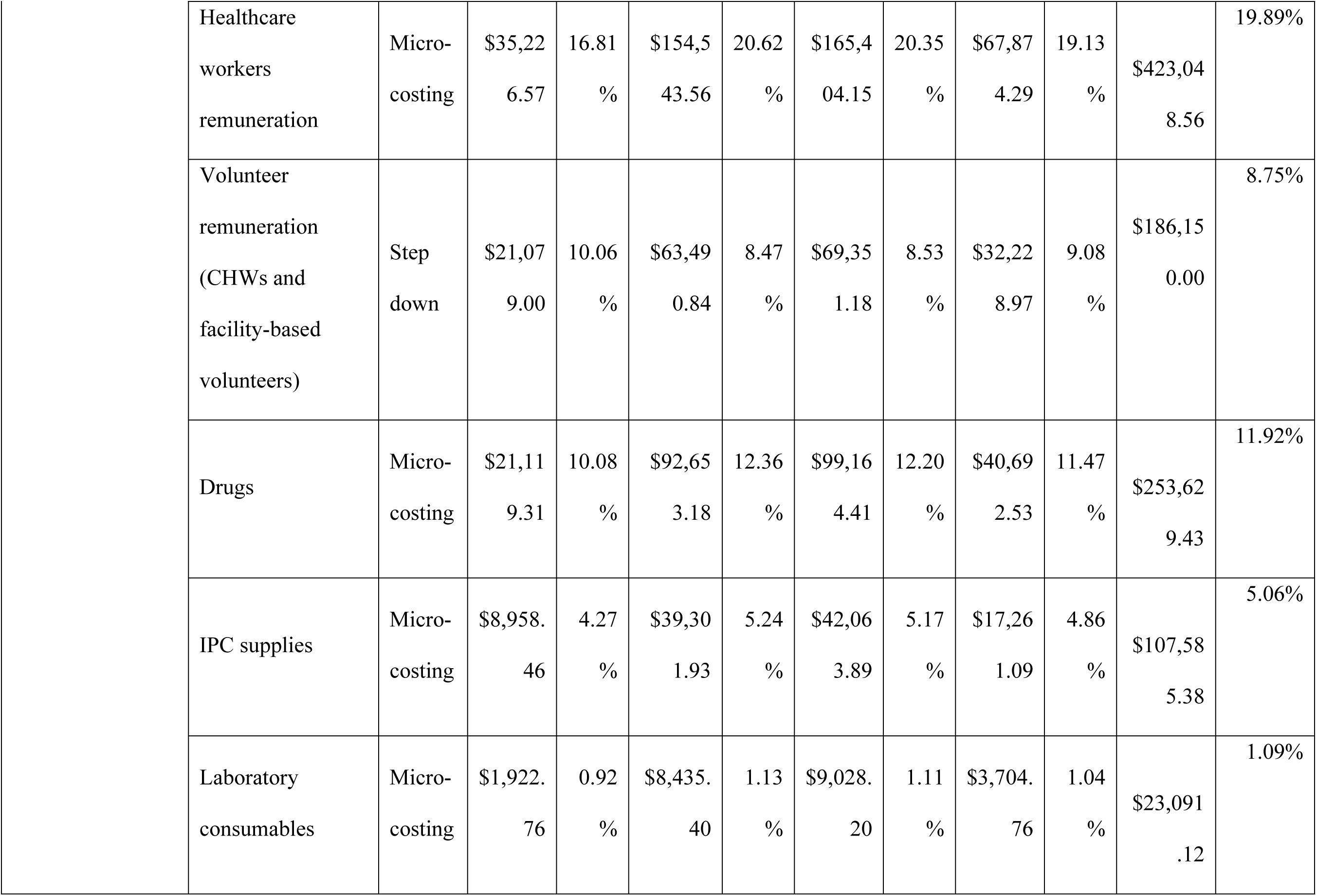
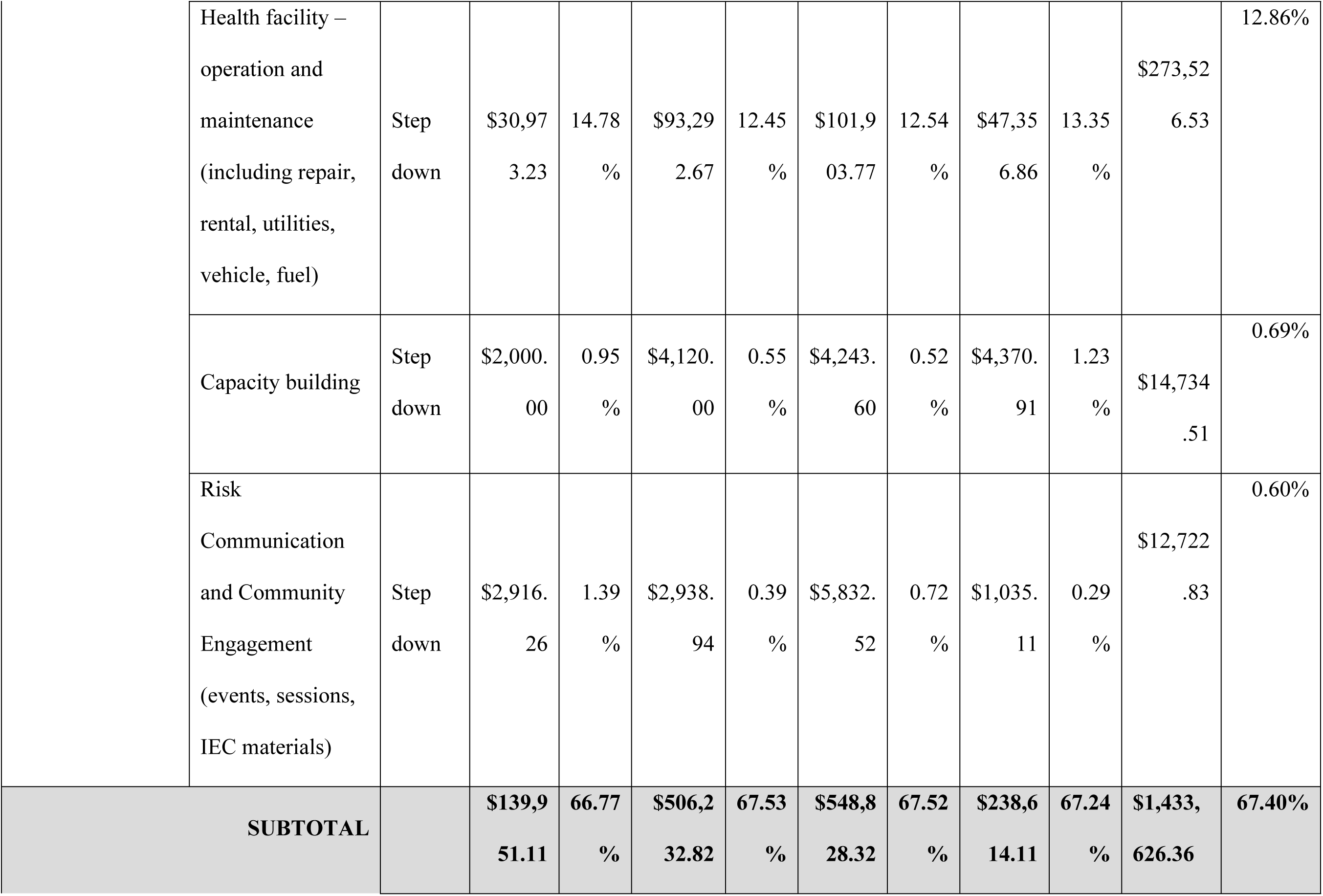
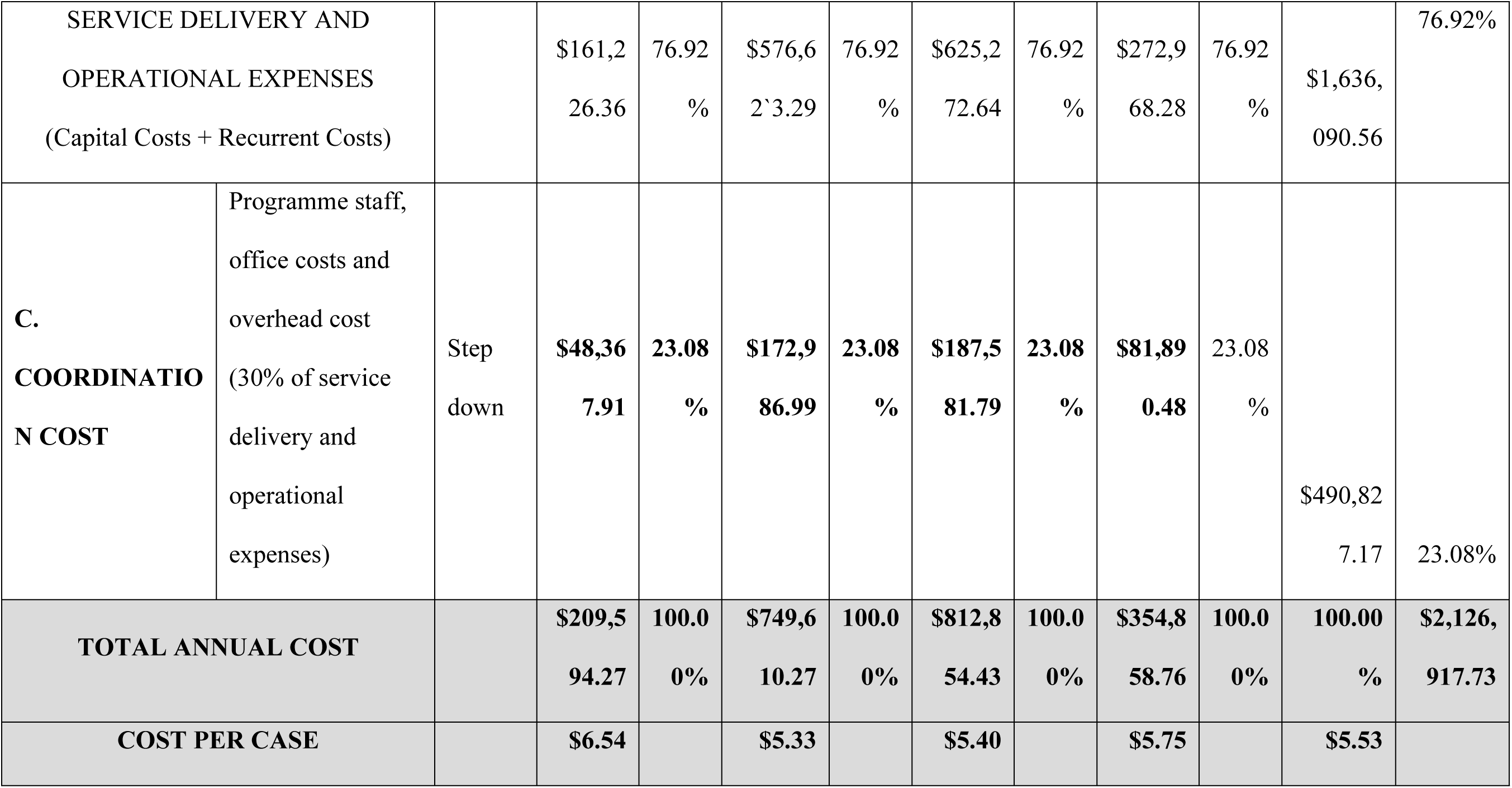
Estimated retrospective cost analysis for scabies outbreak response (2021–2024)

Aligning with the rise of scabies cases from 2021 to 2023, annual cost of scabies-related health service delivery increased four-fold from USD 209,594 in 2021 to USD 812,854 in 2023. Following the MDA campaign, the cost sharply decreased to $354,858.76 in 2024.

The average cost per scabies management ranged between USD 5.33 to USD 6.54, with the highest in 2021 and lowest in 2022.

Healthcare worker remuneration made up 19.89% of the overall cost, consistently sharing the largest portion of the annual cost across the years, ranging from USD $35,226 (in 2021) to $165,404.15 (in 2023). In addition to this cost, $186,150 was required for the remuneration of community health workers and health-facility-based volunteers, which is another 8 – 10% of the annual cost.

The drug costs consisted 11.92% of the overall cost in 4 years. It increased from USD 19,906.98 (11.37%) in 2021 to a peak of USD 99,164.41 (14.08%) in 2023, before dropping to USD 41,913.31 (14.12%) in 2024. The supplementary file 1: details of cost break down provides detail breakdown of the drugs. Of the total cost of $253,629.43 in 4 years, 79% of cost was for permethrin topical cream being used for managing an estimated 85% of the total managed cases. On the other hand, tablet ivermectin only accounted for 2.58% of the drug cost, used for management of an estimated 10% of the caseload.

Additionally, an estimated USD 107,585 was utilized for infection prevention and control and USD 23,091 was utilized for laboratory consumables, making up 5% and 1% of the overall cost in 4 years, respectively. Operational and maintenance expenses for health facilities accounted for an estimated 12 to 15% of the annual cost. This proportion was calculated based on the contribution of scabies to the total number of consultations managed by these facilities.

Investment in risk communication and community engagement and capacity building were relatively low throughout the period, each category sharing less than 1% of the overall annual budget.

**Table 7.** SCENERIO ANALYSIS.

|  | SCENERIO-1 | Ratio | SCENERIO-2 | Ratio | SCENERIO-3 | Ratio |
| --- | --- | --- | --- | --- | --- | --- |
| DRUG COST | \$253,629.43 | 1 | \$211,983.90 | 83.58% | 238,718.36 | 94.12% |
| NON-DRUG COST | \$1,873,288.30 | 1 | \$1,860,794.64 | 99.33% | \$1,868,814.98 | 99.76% |
| TOTAL COST | \$2,126,917.73 | 1 | \$2,072,778.54 | 97.45% | \$2,107,533.34 | 99.09% |

A comparative analysis of different scenarios of proportions of patients treated with topical permethrin and oral Ivermectin.

It demonstrates that existing permethrin-based treatment preference is most expensive treatment modality (Table 1 and Figure 1). Using ivermectin as the preferred treatment (scenario 2) drug costs can be reduced to nearly 83.5% of a permethrin-based approach, saving 2.55% of the overall budget. In case both drugs are utilized in equal proportion (scenario 3), drug costs drop to 94.12% of the current practice with an overall 1% total cost reduction.

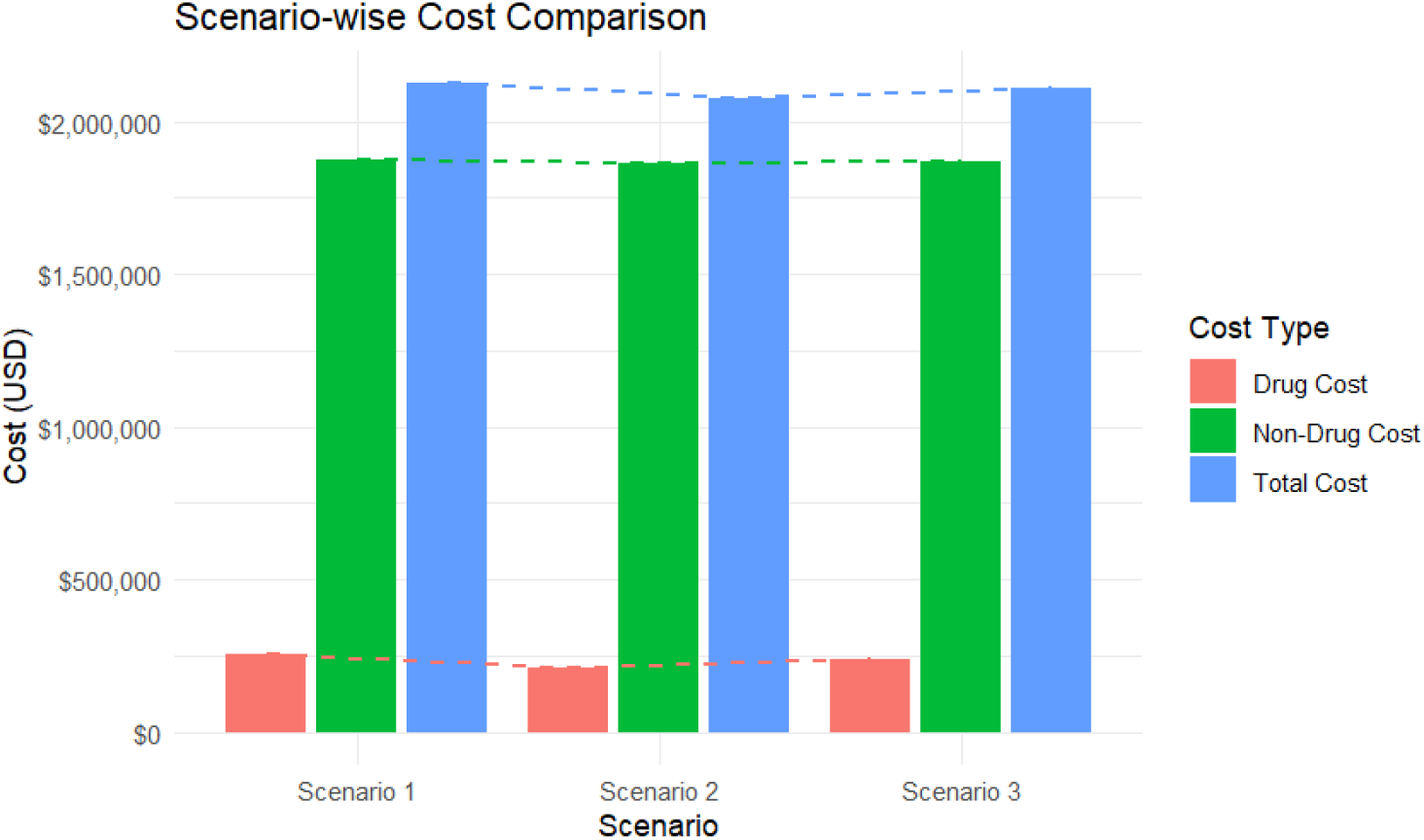

## DISCUSSION

This is the first-ever study in the humanitarian and low-resource setting which explored the economic impact of scabies in health system. The study revealed a high economic impact of scabies on the health system valuing USD 2.12 million over four years with an average treatment cost of USD 5.53 per case. The study also found significant impact of mass drug administration on reducing economic burden on health system.

The average cost per scabies management ranged between USD 5.33 to USD 6.54. Since there was no other cost-analysis study available for scabies at humanitarian crises and massive outbreak setting, we could not compare our results with similar settings. However, the cost in our refugee setting was lower than the hospital setting in India and the primary care setting in Fiji, where the estimated average cost per patient were USD 12.20 and USD 17.7, respectively. The cost per patient also seems lower than other health burden, like tuberculosis, HIV, Hepatitis and non-communicable diseases (4,14). However, if we consider the massive scale of the outbreak, the overall financial cost of the scabies upsurge had significant impact on the health system in the low resource setting.

We have found that the estimated financial cost for the IOM’s scabies outbreak response was USD 2.12 million from 2021 to 2024 with an annual average of USD 531,729. This annual average is 3.33% of the overall funding appeal made by IOM for in health programme in 2024, indicating a significant economic impact of scabies in health response (15). While IOM covers only one-fourth of the health facilities in the refugee camps, extrapolating it to sector wide response may cost around USD 2.1 million for full coverage of the catchment population in a year, which is around 2.5% of the appeal made by the health sector under the Joint Response Plan in 2025 (16). Our finding coincides with the findings of the country-level recent study of Fiji, where the researchers found although a cost measure of single case of scabies is low, extrapolating the finding across the country, the estimated cost of scabies on the Fijian health system was ∼USD 3.0 million per year, equivalent to USD 3.3 per capita (4).

Our study found that annual cost for scabies outbreak response was drastically reduced in 2024 followed by the Mass Drug Vaccination campaign (November 2023 – January 2024), which was 43% of the previous year. Our previous study found that MDA contributed to a significant reduction of scabies cases, which sustained for six months followed by an upward shifting *(Halder et al)*. Aligning with this, our cost-analysis also suggests MDA contributes to significant reduction of economic burden in health systems in high prevalence context. While the estimated cost saving following MDA in IOM facilities was USD 457,996, stimulating it to sector-wide response may lead to a saving of approximately USD 2.0 million with an investment of USD 1M required for the campaign (17). The effectiveness and cost-efficiency of Mass Drug Administration have also been noted in other settings, like Fiji and Ethiopia (8,18). In overcrowded and high burden settings, where the risk factors cannot be addressed properly, by treating the entire community at a time, MDA allows to substantially reduce the scabies prevalence and associated costs. The effectiveness of MDA will only be applicable until the scabies prevalence remains >10% of the community members (18).

We have studied three scenarios to explore the most cost-effective treatment regimen for scabies with first scenario demonstrating the existing practice of permethrin-based treatment, second scenario with ivermectin-based treatment and a third scenario where equal proportion of cases are managed by ivermectin and permethrin. Our analysis found that having ivermectin as preferred treatment (scenario 2) can reduce 16.5% of the drug cost and 2.55% over the overall scabies-related direct and indirect cost. a permethrin-based approach. When both drugs are utilized in equal proportion, 5.88% of drug costs can be reduced, while there will only be a reduction of 1% of the total cost. Topical permethrin and oral ivermectin are both effective and recommended treatment regimen for scabies, although ivermectin has contraindications in case of pregnancy and infants (1,12). A few studies suggest that topical permethrin is clinically more effective than oral ivermectin (6). In India, it was found that although the total cost of treatment with oral ivermectin was lower than topical permethrin, cost for relieving itching and transportation was higher than permethrin (19). In endemic and high-burden setting, ivermectin can be more effective if there remains significant concern on the compliance (18). Therefore, the decision of preferred treatment should not only be decided based on this economic analysis presented here, but also consideration of endemicity, compliance, clinical outcome and response time.

This was a retrospective costing analysis, therefore, in case unavailability of exact data, the researchers had to estimate the nearest possible cost based on the discussion on the relevant stakeholders. Another limitation was our study did not measure the economic impact of scabies from the patient and social perspective. For instance, this massive scabies outbreak may have impact on the quality of life and result in social exclusion and stigma and absenteeism at school and workplace (20). There could have costs of scabies related to productivity loss, time of caregiving and travel. Therefore, while further research is warranted to understand the overall impact of scabies on health system and community, prevention and control strategies, including mass drug administration, should be imposed to reduce the burden of scabies on health system and people’s lives (1).

## CONCLUSION

Scabies has a high impact on health-related quality of life. Our results indicated that, although the average cost of treating scabies is relatively low, overall, the treatment cost for such a large population has a significant economic impact. This study found a substantial effect of mass drug administration on reducing the financial burden on the healthcare system. Strengthening mass drug administration and reinforcing preventive measures is crucial to prevent such an upsurge, especially in low-resourced settings like that of the Rohingya refugee camp.

## DATA AVAILABILITY

All relevant aggregated data and analysis scripts for this article are available from Zenodo at DOI: https://doi.org/10.5281/zenodo.17697474. The dataset includes anonymized, aggregated scabies case counts and climate variables (temperature, rainfall, humidity) for 2021–2023. Related datasets from the same scabies research series are available at DOIs: https://doi.org/10.5281/zenodo.17648621 and https://doi.org/10.5281/zenodo.17687717

## ACKNOWLEDGMENTS

CEH conceptualized and designed the study, provided overall methodological guidance, and led the drafting and critical revision of the manuscript. MAH assisted in refining the study design and drafting the manuscript. SSUZ, JTP, HT supported field coordination, MAH, JA assisted supporting the data collection processes. MAH supported with climatic data triangulation. DWB, ERS and JCO contributed to the critical review and refinement of the manuscript. MFH, ERS, UMP and JCO provided strategic oversight and ensured alignment of the study within broader programmatic and policy frameworks. All authors reviewed and approved the final version of the manuscript. CEH is the corresponding author and JCO is the guarantor of the study.

